# Developmental tuning of prefrontal network fluctuations marks functional maturation in infancy

**DOI:** 10.64898/2026.03.25.26349326

**Authors:** Kanghui Li, Yong Zhang, Yanhui Li

**Author notes:** Corresponding author: Yong Zhang., Yanhui Li.

## Abstract

The early development of the prefrontal cortex is crucial for higher cognitive functions. However, current research presents inconsistent findings regarding whether intra-prefrontal connectivity increases or decreases in infants younger than six months. Do dynamic changes in connection strength across different states over time carry information about prefrontal maturation? This study used functional near-infrared spectroscopy (fNIRS) to record prefrontal brain activity in 48 healthy infants aged 1-8 months during natural sleep and auditory stimulation. By analyzing the fluctuations in frequency-domain characteristics of functional connectivity (FC) and various brain network properties, we found that: under auditory stimulation, the intensity of FC fluctuations in the ultra-low frequency range was positively correlated with age; while in the resting state, the fluctuation intensity of network properties in relatively higher frequency bands decreased with age. Furthermore, auditory stimulation reconfigured the energy distribution of network fluctuations, shifting it towards higher frequency bands. These results suggest that the early development of the infant prefrontal internal network is characterized by state-dependent optimization of its dynamic fluctuation properties, shedding light on the developmental tuning of functional network dynamics in infancy.

## 1. Introduction

The prefrontal cortex of the human brain is an extremely important and complex region, often referred to as the brain’s "command center" (Miller EK, 2000; Friedman NP et al., 2022). It is the core brain region responsible for critical human abilities such as intelligence (Cole MW et al., 2015), memory (Funahashi S et al., 2006), and attention (Bahmani Z et al., 2019). The development of the prefrontal cortex begins in the fetal stage (Desrosiers J et al., 2024) and continues after birth (Cao M et al., 2017; Gao W et al., 2011). Both the thickness and surface area of the gray matter change as a person ages (Zhang H et al., 2022). However, compared to other areas of the cerebral cortex, it is considered to mature significantly later (Casey BJ et al., 2000; Dennis EL et al., 2013). At birth, the auditory and visual cortices already exhibit adult-like connectivity patterns (Hoff GE et al., 2013), but the prefrontal cortical network begins to resemble adult-like connectivity only around the age of 1 (Gao W et al., 2009), and reaches approximately 80% of adult connectivity by the age of 4 (Sun L et al., 2025). During the earliest postnatal period, specifically between 0 and 6 months, the functional connectivity of the prefrontal cortex with other brain regions, especially long-range connections, has been found to increase (Homae F et al., 2010). However, current research on the network development within the prefrontal cortex remains insufficient, with a wide range of age spans (Kardan O et al., 2022; Cao M et al., 2017). Due to the large age range, we are unable to determine what changes occur within the prefrontal cortex during the critical early period of 0-6 months. The few existing studies on the connectivity within the prefrontal cortex show contradictory results, with some indicating enhancement and others suggesting a decrease (Homae F et al., 2010; Gao W et al., 2015; Yin W et al., 2019). However, during the first six months of life, cognitive development occurs at such a rapid pace (Hua J et al., 2019). For instance, a 3-month-old infant shows a desire to reach for an object but cannot yet control their hand to grasp it. By 5 months, most infants are able to effectively use their hands to grab objects they see. The responses to external stimuli and methods of self-entertainment differ significantly between 1-month, 3-month, and 6-month-old infants. We argue that relying solely on overall connectivity strength omits too much developmental information. We believe that hidden within the constantly changing network fluctuations of the brain are more insights into infant brain development.

In this study, we record prefrontal cortex activity in infants using near-infrared functional neuroimaging (fNIRS) under two different brain states: auditory stimulation and resting state. We ask: Is there sensitive developmental evidence of the internal network within the prefrontal cortex during the first six months after birth? Our three main objectives are: First, we aim to determine whether the fluctuations in functional connectivity (FC) and various brain network properties change with age, and which network metric is most sensitive to age-related fluctuations. Second, we aim to explore the differences in fluctuations between the resting state and the auditory stimulation state. Finally, we investigate whether the characteristic frequency domains change with age or state. (The classic definition of resting state refers to spontaneous fluctuations while awake without any specific task. For convenience in this study, we refer to the natural sleep state without auditory stimulation as "resting state.")

## 2. Methods

### 2.1. Participants

The experiments conformed to the Declaration of Helsinki, and the experimental protocol was approved by the Ethics Committee of the Xi’an People’s Hospital (Xi’an Fourth Hospital) (ethical approval number was KJLL-Z-K-2024047). Written informed consent was obtained from the parents or legal guardians of all infant participants prior to the study. This study is a retrospective study that utilized anonymized clinical data from December 2023 to December 2024. All patient information has been removed to protect patient privacy. This study included a total of *n* = 48 full-term healthy infants (mean age, 108 days; range, 30-260 days; 25 males), of which *n* = 22 received auditory stimulation (mean age, 88 days; range, 30-235 days; 9 males). We divided the *n*=48 participants into three groups based on their age in months: <3m (*n*=19), 3-4m (*n*=14), and >5m (*n*=15). The *n*=22 participants who received stimulation were divided into three groups based on their age in days: <60d (*n*=8), 60-100d (*n*=8), and >100d (*n*=6). All participants passed a hearing test, and all infants underwent developmental assessments by pediatricians to ensure there were no developmental delays or issues with muscle tone.

### 2.2. Stimuli

We selected white noise, the most ’meaningless’ sound with the most uniform spectrum, as the auditory stimulus. Unlike human voices, music, or other stimuli, white noise carries no linguistic or emotional information, making it an ideal pure auditory stimulus. This allows us to effectively rule out interference from higher-level cognitive processes such as language processing or emotional responses. Previous study has confirmed that white noise stimulation indeed elicits hemodynamic responses in the prefrontal cortex of sleeping infants (Li K et al., 2025). All infants underwent fNIRS recordings after entering natural sleep. First, a 5-minute resting-state signal was collected, followed by exposure to the auditory stimulus. The original audio files can be found in previous study (Li K et al., 2025). The auditory stimulation consisted of 15 seconds of sound followed by 20 seconds of silence, repeated for five cycles (Fig. 1e). The distance between the sound source and the top of the infant’s head was approximately 75 cm, with a sound intensity of 54.7 dBA. During the entire detection process, parents sat comfortably in a chair with their arms still, while the infants’ ears were fully exposed.

**Fig. 1.**
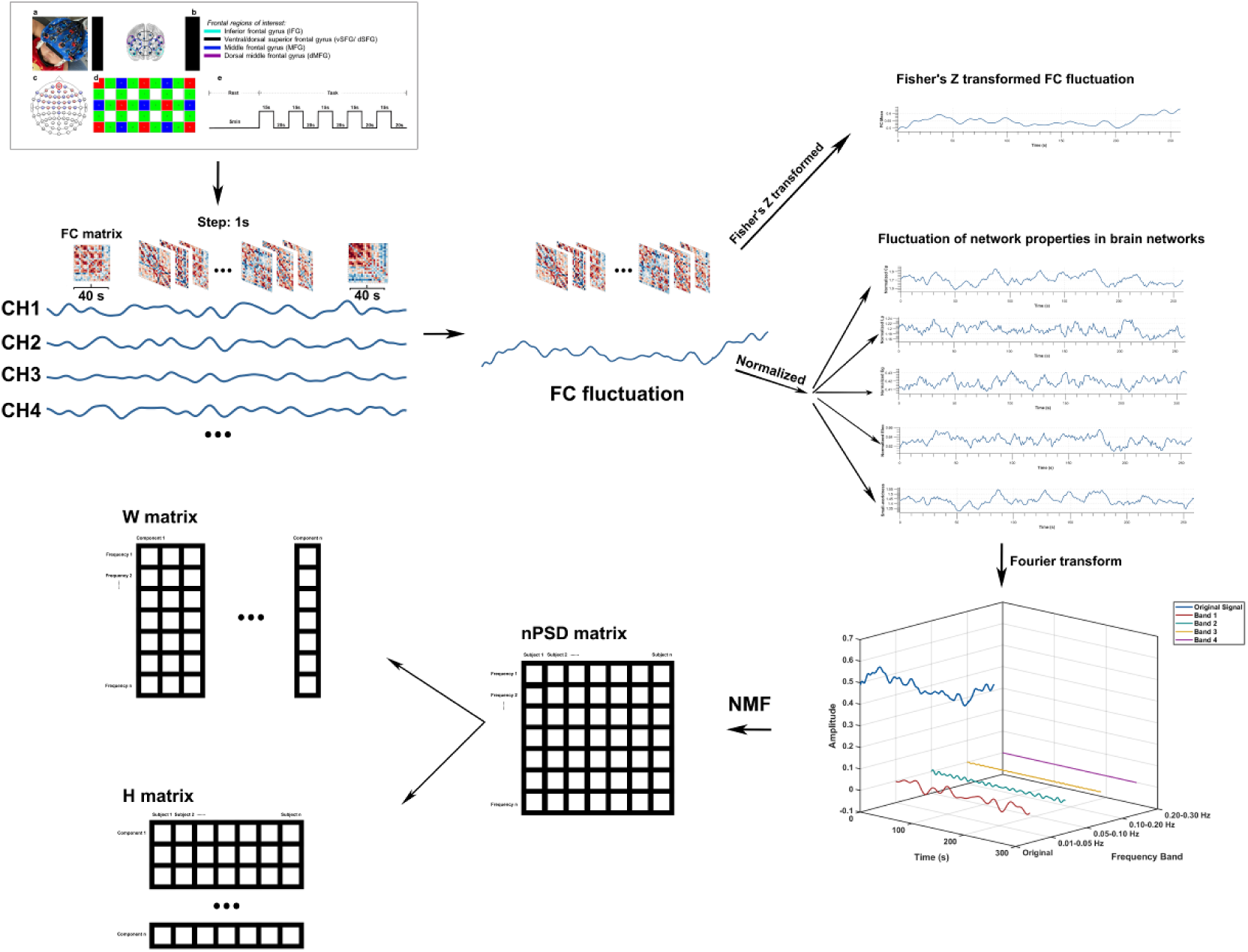
**∣ Analysis of fluctuations in prefrontal cortex functional connectivity and network Properties**. **a**,The probe set was placed on the frontal in accordance with the 10-10 EEG system. **b**,Channel locations correspond to brain regions as referenced in Laurie Bayet’s article (Bayet L et al., 2021). **c**,The 17th source probe is located at the FPz site (red circle in the image). **d**,A total of 15 probes was used, with the green ones representing the 22 channels. **e**,The sound-stimulation procedure included 5 minutes of resting-state data collection during natural sleep, followed by sound stimulation consisting of 15 seconds of stimulation and 20 seconds of resting time, repeated for a total of 5 cycles. By preprocessing the raw data collected, functional connectivity (FC) strength within 40-second sliding windows was computed for 22 channels covering the prefrontal cortex, with a sliding step of 1 second. This generated FC matrices that varied over time. Subsequently, the fluctuation curves of FC and network properties were derived by sequentially calculating these matrices. The fluctuations were then subjected to Fourier transformation, followed by feature extraction of the frequency domain using Non-negative Matrix Factorization (NMF).

### 2.3. fNIRS Data acquisition and preprocessing

fNIRS signals were collected using a multichannel fNIRS system (ETG-one, Hitachi) operating at two wavelengths (695 and 830 nm) with a sampling frequency of 10 Hz. The setup included 8 source probes and 7 receiving probes, spaced 3 cm apart (Fig. 1d). Probe placement followed the international 10-10 EEG positioning system, with the 17th source probe positioned at the FPz location (Fig. 1c). The channels’ locations correspond to specific brain regions as outlined in Laurie Bayet’s study (Bayet L et al., 2021) (Fig. 1b).

We first preprocessed the raw data using the NIRS_KIT software (Hou X et al., 2021) (version 3.0_Beta, https://github.com/NIRS-KIT), based on Matlab (version 2023b, https://www.mathworks.com/). A polynomial regression model was applied for detrending to estimate the linear trend, while motion correction was carried out using the Temporal Derivative Distribution Repair (TDDR) method (Fishburn FA et al., 2019). To minimize the impact of high-frequency systemic physiological noise, such as respiration and heartbeat, along with artifacts due to very low-frequency drift, a band-pass filter with an Infinite Impulse Response (IIR) was used, targeting the 0.01–0.08 Hz frequency range.

### 2.4. Data analyses

#### 2.4.1. Calculation of fluctuating network properties

We used the sliding window method for dynamic functional connectivity analysis. The sliding window length was set to 40 seconds, with overlapping windows and a step size of 1 second. The Pearson correlation coefficient between channels within each window was calculated, forming a dynamically changing FC matrix. The Fisher’ s Z transformation was applied to the FC matrix to calculate the average FC for each matrix, thus obtaining the curve of FC fluctuations over time.

The calculation of network properties was based on the FC matrix of each sliding window. The computation of brain network properties was conducted using the software FC_NIRS (version 3.0, https://github.com/FC-NIRS/FC-NIRS) (Xu J et al., 2015). For network properties, we calculated both the real network and the random network, with the random network based on 1000 random calculations. The network type was based on weighted networks, and network density was controlled by calculating Pearson correlation coefficients at 30% sparsity and ≥0.6. The reason for selecting a Pearson correlation coefficient ≥0.6 is that it is widely considered to have a significant correlation in connectivity properties. The choice of 30% sparsity is based on previous research (Li K et al., 2025) as well as studies by Nazari R (Nazari R et al., 2023), which showed that a sparsity of 30% can effectively differentiate between different groups. Nazari R found that, when calculating the correlation between network properties and age, 30% and 40% sparsity adequately reflect the significance of different brain regions, while excessively high or low sparsity is not effective. We calculated the following network properties:

The clustering coefficient (*C*_p_) (Onnela JP et al., 2005) is used to quantify the tendency of network nodes to cluster. For node 𝑖, its clustering coefficient represents the ratio of the actual number of edges between its neighboring nodes to the total number of possible edges, reflecting the prevalence of cascading triangular structures. For an undirected weighted network, the local weighted clustering coefficient of node 𝐶_𝑖_ is defined as:

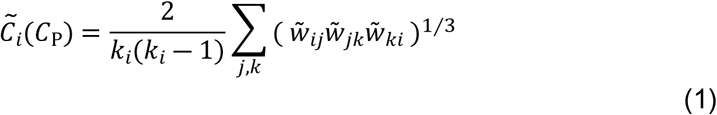

Where 𝑘_𝑖_ is the degree of node 𝑖, and 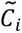 is the normalized edge weight, which scales all weights to the range [0,1]. The sum is taken over all neighboring node pairs (*j*, *k*)of node 𝑖.

The shortest path length (*L*_p_) (Onnela JP et al., 2005) is a global metric used to measure the average path length for information transmission or interaction between all pairs of nodes in a network. A smaller value indicates better connectivity between nodes in the network and higher information transmission efficiency. 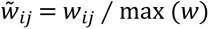 is defined as the shortest path with the strongest weight among all paths connecting nodes 𝑖 and 𝑗, and its mathematical expression is:

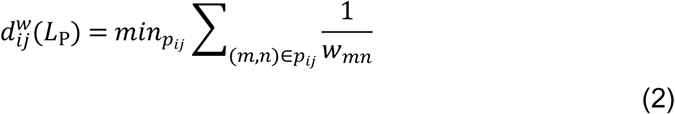

Where 𝑝_𝑖𝑗_ represents a path connecting nodes 𝑖 and 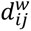 is an edge on the path 𝑝_𝑖𝑗_ . 𝑤_𝑚𝑛_ is the original weight of edge (𝑚,𝑛), and 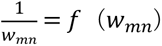 is a function that maps the original weight to a value used for calculating the path length or cost.

Global efficiency (Eg) (Latora V et al., 2001; Wang J et al., 2015) is an important metric used to quantify the overall efficiency of information transmission in a network. The core concept is to treat a network as an information transmission system, measuring the average difficulty of communication between any two nodes in the system. A higher value indicates greater average communication efficiency between network nodes and stronger ability for rapid information transmission. Its mathematical expression is as follows:

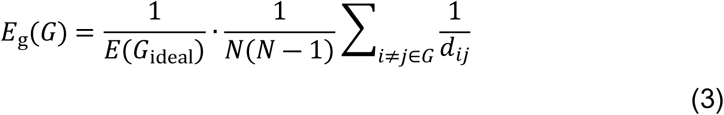

Where 𝑑_𝑖𝑗_ is the shortest weighted path length between nodes𝑖and𝑗, and 𝐸(𝐺_ideal_) is the efficiency of the ideal fully connected graph, which is defined as: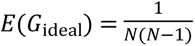 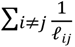 where 𝓁_𝑖𝑗_ is the physical distance or edge weight between nodes 𝑖 and 𝑗.

Local efficiency (*E*_loc_) (Latora V et al., 2001; Wang J et al., 2015) is an important metric used to quantify the local information transmission efficiency and fault tolerance of a network. The core concept is to measure the information transmission efficiency of the subnetwork formed by a node’s direct neighbors from the local perspective of the node. A higher value indicates that alternative communication paths between neighboring nodes are still high, ensuring the stability and robustness of the local network. Its mathematical expression is as follows:

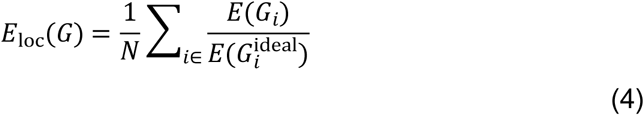

Where 𝐺_𝑖_ is the subgraph formed by all the neighbors of node 𝑖, excluding 𝑖 itself, 𝐸(𝐺_𝑖_) is the global efficiency of the subgraph 𝐺_𝑖_, and 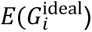 is the efficiency of the ideal fully connected graph corresponding to the subgraph 𝐺_𝑖_.

Small-worldness [40] is used to describe a special network structure that possesses both a high clustering coefficient and a short average path length, indicating global efficient information transmission. It is neither completely regular nor completely random, but exists in an intermediate state, achieving functional optimization. The mathematical expression for the small-worldness scaling coefficient 𝜎 is as follows:

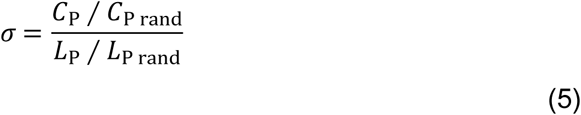

Where 𝐶_P_ is the clustering coefficient of the real network, 𝐶_P rand_ is the clustering coefficient of the random network, 𝐿_P_ is the average shortest path length of the real network, and 𝐿_P rand_ is the average shortest path length of the random network.

#### 2.4.2 Fourier transform

The Fourier transform is an important mathematical tool used to convert a signal from the time domain to the frequency domain. It works by representing complex signals, continuous or discrete in time, as a linear combination of sine and cosine waves with different frequencies, amplitudes, and phases. Through the Fourier transform, a time-domain signal can be decomposed into its constituent frequency components, thereby revealing the frequency structure and energy distribution characteristics of the signal. We perform the Fourier transform of all network fluctuations to obtain the power spectral density (PSD) of all fluctuations using the Welch method. The mathematical expression is as follows:

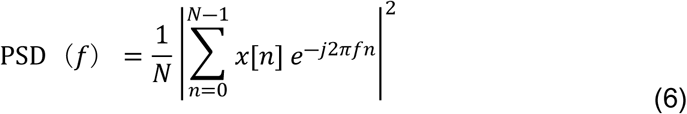

Where 𝑓 is the frequency, 𝑁 is the number of data points, and 𝑥[𝑛] is the time series data of network oscillations. The signal is windowed using the Hamming window to reduce spectral leakage. To make the computed PSD comparable across individuals, interpolation is used to calculate the PSD for every 0.001 Hz. Specifically, the PSD value is calculated for each frequency point, and then linear interpolation is applied to compute the PSD values at every 0.001 Hz, thereby aligning the frequency range for easier comparison and obtaining a finer-grained frequency distribution.

#### 2.4.3. Calculation of nPSD

To facilitate comparison between different subjects, as well as for subsequent non-negative matrix factorization and Shannon entropy calculations, we normalized the PSD for each subject and represented it as the normalized power spectral density (nPSD), which reflects the relative energy contribution. The mathematical expression is as follows:

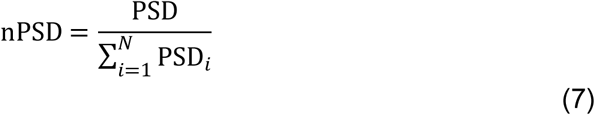

Where PSD represents the raw power spectral density data, PSD_𝑖_is the power value at the 𝑖-th frequency point, 𝑁 is the total number of frequency points, and 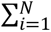 PSD_𝑖_ is the total power spectral density across all frequency points, i.e., the total area under the curve (AUC), which is used for normalization.

#### 2.4.4. Shannon entropy calculation based on nPSD

Shannon entropy is a measure used to quantify the uncertainty or the informational complexity of a variable. When applied to the signal domain, analyzing the signal’s PSD leads to the derivation of Spectral Entropy. To convert the PSD into an effective probability mass function, the PSD is normalized according to equation (7), ensuring that the sum of the probability distribution equals 1. The higher the calculated entropy value, the flatter the nPSD distribution, meaning that the signal power is broadly and uniformly distributed across frequencies. A lower entropy value indicates that the nPSD distribution is more concentrated, meaning the signal power is highly concentrated at one or a few frequencies. Assume that nPSD is the normalized PSD, where the value at each frequency point represents the power proportion of that frequency component, satisfying the following conditions:

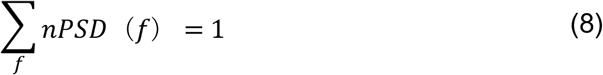

The mathematical expression for the Shannon entropy 𝐻_nPSD_ of nPSD is as follows:

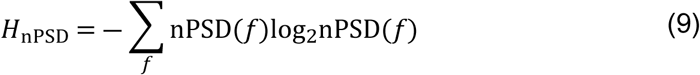

Where nPSD(𝑓) represents the normalized power spectral density at frequency 𝑓, which is equivalent to the probability of that frequency component. The use of log_2_ is because Shannon entropy measures the amount of information, which is the average uncertainty of unit information. The unit is typically in bits, i.e., based on binary computation.

#### 2.4.5. NMF

Non-negative Matrix Factorization (NMF) is an unsupervised technique for linear dimensionality reduction and feature extraction. Given a matrix V(*m×n*) where all elements are non-negative, the goal of NMF is to find two non-negative matrices W(*m×k*) and H(*k×n*) such that their product approximates the original matrix. We first use the elbow method to determine the number of features 𝐾, and then apply NMF to the nPSD. The mathematical expression is as follows:

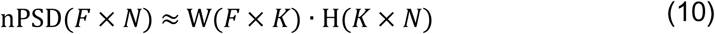

Where nPSD is the matrix composed of the nPSD values for all participants, with dimensions of 𝐹 frequency points by 𝑁 participants. W is the basis matrix with dimensions of 𝐹 frequency points by 𝐾 components, where each column represents an extracted feature frequency distribution. H is the coefficient matrix with dimensions of 𝐾 components by 𝑁 participants, where each row represents the weight of each participant in the feature frequency domain.

#### 2.4.6. General linear mixed model (GLMM) for the age-related analysis of PSD curve AUC

We established the following Generalized Linear Mixed Model (GLMM) to examine the relationship between the area under the curve (AUC) of power in a certain frequency band and factors such as age:

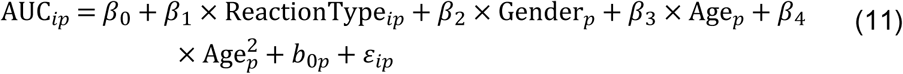

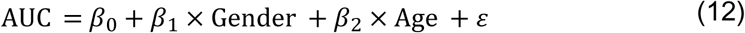

Formula (11) represents the Generalized Linear Mixed Model (GLMM) for examining the correlation between FC fluctuation PSD AUC and age. In this model, AUC_𝑖𝑝_denotes the area under the curve (AUC) for the 𝑖-th response type of the 𝑝-th participant. 𝛽_0_ is the fixed effect intercept, 𝛽_1_ is the fixed effect coefficient for response type, 𝛽_2_ is the fixed effect coefficient for gender, and 𝛽_3_ and 𝛽_4_ are the fixed effect coefficients for age and its squared term, respectively. The inclusion of the squared term aims to detect any potential non-linear relationship between age and AUC. 𝑏_0𝑝_ is the random intercept for the 𝑝-th participant included in the model, which follows a normal distribution with a mean of zero. This random effect is based on the response interaction terms and is intended to control for variation caused by within-group correlations. 𝜀_𝑖𝑝_is the residual term of the model, which also follows a normal distribution with a mean of zero.

Formula (12) represents the General Linear Model (GLM) for examining the correlation between the fluctuation PSD AUC of all network properties and age. 𝛽_1_is the fixed effect coefficient for gender, 𝛽_2_ is the fixed effect coefficient for age, and ɛɛ is the residual term of the model.

#### 2.4.7. The calculation of nSNR and nDR

To eliminate the dimensional differences between Signal-to-Noise Ratio (SNR) and Dynamic Range (DR) and allow for comparison on the same scale, we performed Min-Max Normalization on their group mean and the standard error of the mean, so that their mean values are distributed within the [0,1] range. The normalization process for any given metric (SNR or DR) is as follows:

Mean normalization:

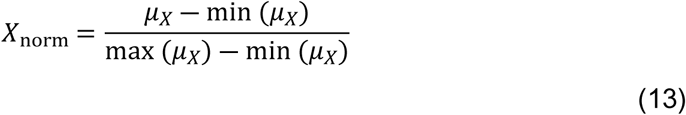

Standard error normalization:

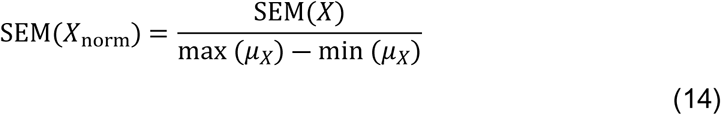

Where 𝑋_norm_ represents the normalized mean of nSNR and nDR, 𝜇_𝑋_ represents the mean vector of the original metrics calculated over different window lengths, min (𝜇_𝑋_) and max (𝜇_𝑋_) represent the minimum and maximum values in the mean vector, respectively, SEM(𝑋) represents the original standard error, and SEM(𝑋_norm_) represents the normalized standard error.

All network properties were standardized before the analysis, as shown in the following formula:

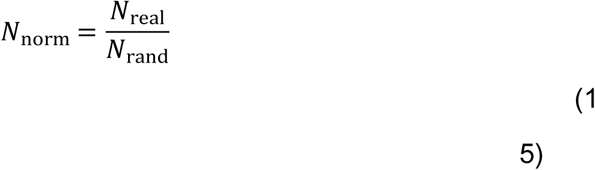

Where 𝑁_norm_ is the standardized network properties, 𝑁_real_ is the real network properties, and 𝑁_rand_ is the random network based on 1000 random calculations.

## 3. Results

In this paper, we collected resting-state fNIRS data from the prefrontal cortex of 48 healthy full-term infants aged 1 to 8 months. We successfully completed auditory stimulation in 22 cases (fig. 1e). In the natural sleep state, we analyzed the intra-prefrontal cortex functional connectivity strength and fluctuations of different network properties under two different states in infants, and used this time-varying information to search for evidence of the development of the prefrontal cortex internal network. The infants who completed auditory stimulation were selected from the 48 in the resting state.

We first conducted the selection of the optimal sliding window. For the resting-state FC of *n*=48 infants, we analyzed simple fluctuation state attributes such as the mean, variance, and standard deviation of fluctuations across different age groups and correlations (fig. 2a). We performed a Fourier transform on the FC fluctuations in the resting state to extract the power spectral density (PSD) (fig. 2b). The PSD was standardized (Methods) and, through non-negative matrix factorization (NMF) (Methods) (Tang X et al., 2021), we extracted feature frequencies (fig. 2d).

**Fig. 2.**
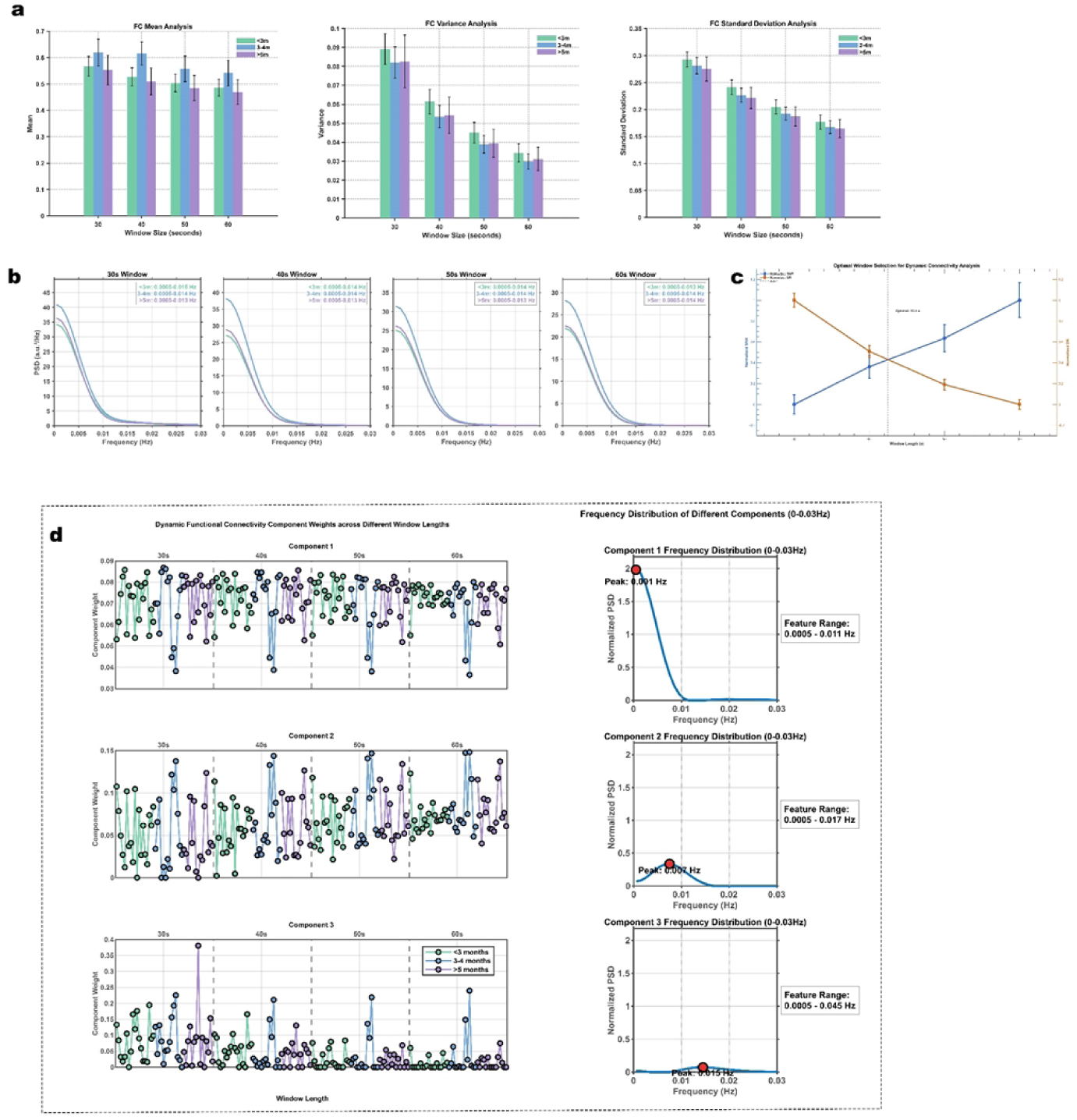
**∣ Evaluation of Different Window Lengths**. **a**,The mean, variance, and standard deviation of FC fluctuations in different age groups were assessed using various window lengths (30s, 40s, 50s, and 60s). **b**,Power Spectral Density (PSD) of FC fluctuations in different age groups was evaluated across different window lengths. **c**,The normalized Signal-to-Noise Ratio (nSNR) and normalized Dynamic Range (nDR) of FC fluctuations were computed for all subjects (*n*=48) across different window lengths to identify the optimal window length. **d**,The normalized PSD (nPSD) for different window lengths (*n*=192) were subjected to Non-negative Matrix Factorization (NMF), and the characteristic frequency domains (right) and their associated frequency domain weights (left) were compared across different age groups.

The purpose of using different age groups was very important: to prove that the fluctuations detected in FC using the sliding window method were truly dominated by the real functional connectivity fluctuations of the prefrontal cortex, not by noise generated during the sliding process. This was evident regardless of the window size used, as we found consistent distribution patterns across the three age groups (fig. 2a,b,d), whether for basic metrics like the mean or for frequency-domain decompositions such as the PSD. To find the optimal window length, we used the balance point of signal-to-noise ratio (SNR) and dynamic range (DR).

SNR is an indicator of fluctuation quality, and its higher value means the signal is clearer with less noise pollution (Stam L et al., 2019). DR measures the ratio between the ability to process the strongest and weakest signals, and its higher value means the ability to detect a wider range of fluctuation changes (Titze IR et al., 2017). Longer window lengths increase the SNR but reduce the DR, which means longer windows reduce the noise from the sliding window but also lose information about the real FC fluctuations. Therefore, we selected the balanced point of these two metrics as the criterion for choosing the most appropriate window length.

Since SNR and DR are two physical quantities with different dimensions and value ranges, it is meaningless to directly compare them on one graph. Therefore, we performed Min-Max Normalization (Methods) on both SNR and DR to find their intersection. As shown in fig. 2c, the intersection of nSNR and nDR occurs at 42.5s. Combining this with fig. 2d, we observed that the relative position of component weights began to stabilize after 40s. Therefore, we selected 40s as the most suitable sliding window length.

### 3.1. The internal network fluctuations of the prefrontal cortex in infants aged 1 to 8 months change with age

We performed an analysis of prefrontal cortex (FC) fluctuations and found that age effects were only observable in the auditory stimulation state with *n*=22 infants. Due to the large inter-group sample differences caused by grouping individuals based on their chronological age, we grouped them according to their exact age in days (Methods). In the resting state, the PSD curve shapes of different age groups showed little difference (fig. 3c), whereas in the auditory stimulation state, the PSD curves of different age groups displayed significant differences (fig. 3d)( ANOVA ; F(2, 19) = 10.20, *p* = .001, *ω*² = 0.46, 95% CI [0.043, 0.634]), primarily in the ultra-low-frequency range. We used NMF to extract the characteristic frequency domain range of FC fluctuations within 0-0.1 Hz. Standardized power spectral density (nPSD) (Methods) was subjected to NMF, which yielded the first principal component at 0.0005-0.011 Hz (fig. 3h). We compared the area under the PSD curve (AUC) of Principal Component 1 across different age groups (fig. 3f) and observed significant inter-group differences (F(2, 19) = 10.20, *p* = 0.001, *ω* ² = 0.46, 95% CI [0.043, 0.634]). No significant inter-group differences were observed in the resting state (fig. 3e).

**Fig. 3.**
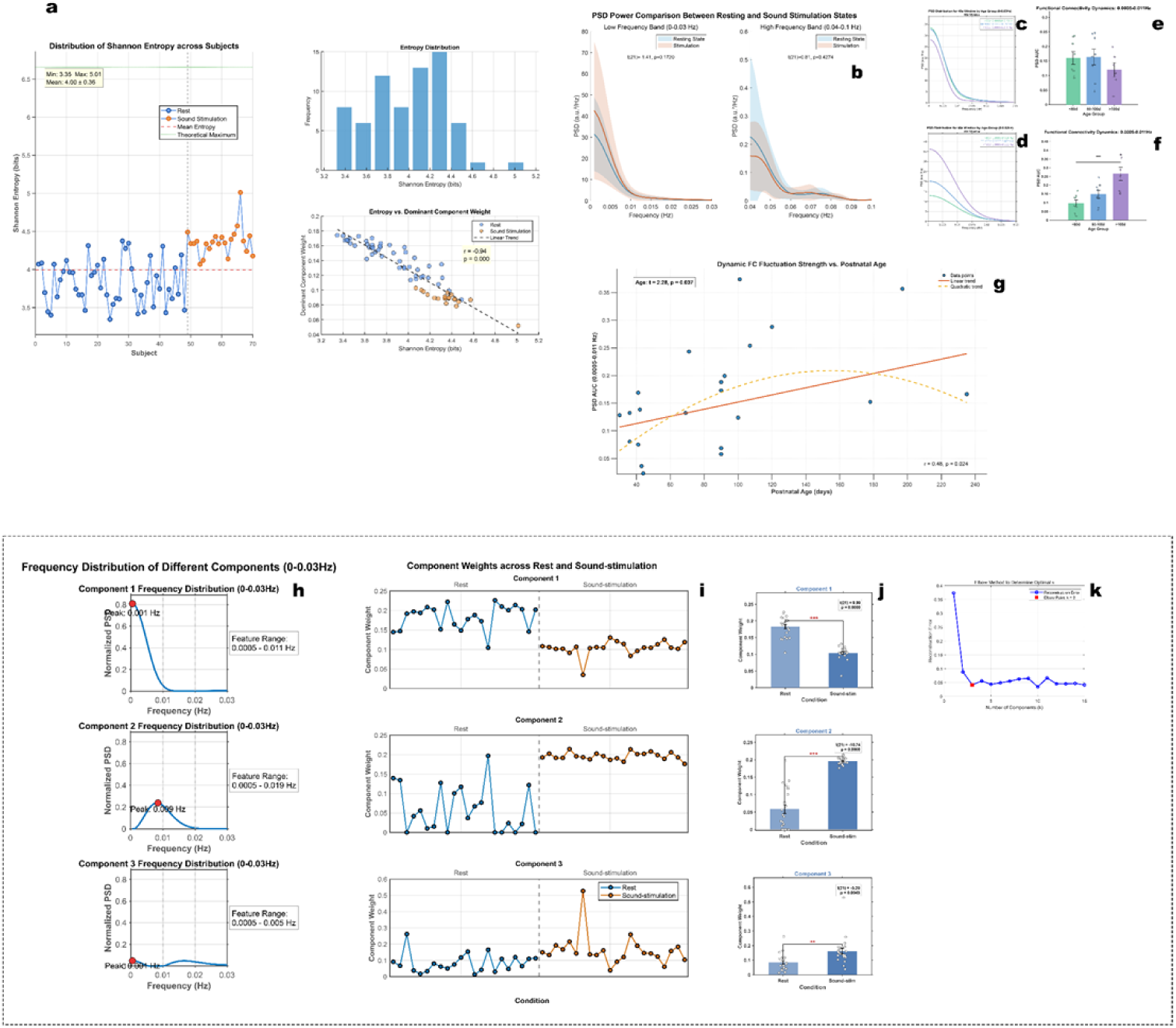
**∣ The fluctuation of functional connectivity (FC) in the prefrontal cortex**. **a**,Normalized Power Spectral Density (nPSD) complexity analysis of FC fluctuations in infants (*n*_rest_=48,*n*_stimuli_=22) during natural sleep after sound stimulation. The left figure shows the Shannon entropy of nPSD for each individual, while the right figure represents the distribution of Shannon entropy and the relationship between entropy and the weight of the dominant components. **b**,Comparison of resting and sound stimulation PSD AUC in the 0-0.03 Hz and 0.04-0.1 Hz frequency ranges (*n*=22). **c**,Shape of the PSD curve during rest (*n*=22) in different age groups. **d**,Shape of the PSD curve during sound stimulation (*n*=22) in different age groups. **e**,The frequency range (0.0005-0.011 Hz) of component 1 extracted by NMF. Comparison of the AUC of resting-state PSD (*n*=22) between different age groups. **f**,Comparison of AUC of sound stimulation PSD (*n*=22) in the 0.0005-0.011 Hz range between different age groups (ANOVA; *p* < 0.001;*ω*² = 0.46; 95% CI,(0.04, 0.63)). **g**,Generalized Linear Mixed Model (GLMM) of the AUC of PSD and age in the 0.0005-0.011 Hz range under sound stimulation. The solid line represents Age (*p* = 0.037;*β* = 0.0028;95% CI(0.0002, 0.0055)), and the dashed line represents Age² (*p*=0.083) Pearson correlation analysis (bottom right; *r*=0.48; *p*=0.024). **h**, NMF decomposition of nPSD for both resting and sound stimulation (*n*=44) to extract 3 characteristic frequency domains. **I**,Comparison of the weight of each characteristic frequency domain in resting and sound stimulation states. **j**,Paired *t*-test of component weights before and after stimulation. **k**,Confirmation of the optimal K value for NMF using the elbow method. (*\*p* < 0.01; *\*\*p* < 0.01; *\*\*\*p* < 0.001)

Since previous studies have confirmed that infants show different FC response patterns to auditory stimuli (Li K et al., 2025), including Sensitive-Positive, Sensitive-Negative, and Insensitive responses, we analyzed a Generalized Linear Mixed Model (GLMM) that included response type, gender, age, and the nonlinear dynamics of age (fig. 3g). The main effect of age showed a significant linear growth trend (F(1, 16) = 5.20, *p* = 0.037; *β* = 0.0028, 95% CI [0.0002, 0.0055]). The trend test for the quadratic term of age did not reach significance (F(1, 16) = 3.42, *p* = 0.083), but it was close to being marginally significant, suggesting that this growth effect has some nonlinear dynamics, although the nonlinearity might not be significant due to the small sample size. The main effects of response type and gender were not significant. We also performed an age-related analysis of the second principal component of NMF (0.0005-0.019 Hz) (Supplementary Fig. 1), and the results remained significant, although less pronounced than for the first principal component. We conducted the same PSD AUC analysis on the entire resting-state data of *n*=48, but no age-related frequency domain features were observed.

The network properties based on graph theory represent the connectivity between different elements (Koutrouli M et al., 2020). Each brain region is defined as a node, and the connection between brain regions is defined as an edge. This network of edges and nodes forms the brain network, which is an efficient information processing architecture evolved through natural selection to achieve the optimal balance between functional specialization and global information integration (Thiebaut de Schotten M et al., 2022). It is necessary to support the complex dynamic processes required for generating intelligence, consciousness, and thought (Chen H et al., 2023; Malagurski B et al., 2019). The fundamental language rules followed by these network properties are optimization and trade-offs (Thiebaut de Schotten M et al., 2022), self-organized criticality (Racz FS et al., 2018), and dynamic adaptation.

In the resting state (*n*=48), we observed age-related changes in the fluctuations of network properties in relatively higher frequency domains. Unlike FC fluctuations in task states, the intensity of these fluctuations in the frequency domain decreased with age. We first performed NMF decomposition on the nPSD within 0-0.1 Hz. Due to the large magnitude difference between the ultra-low frequency domain and relatively higher frequency domains, the characteristics of the higher frequencies were completely masked. Therefore, we performed a separate NMF analysis on the frequency domain above 0.04 Hz and found that many characteristic frequency domains were separated in the 0.04-0.1 Hz range. Although the elbow method suggested K=5, a clear downward trend was still observed later (fig. 4h). Therefore, the separated features may be overly complex, making it difficult to identify the primary frequency domain range. We subsequently divided the 0-0.1 Hz range into five non-overlapping frequency domains to discover the main age-related characteristic frequency domains.

**Fig. 4.**
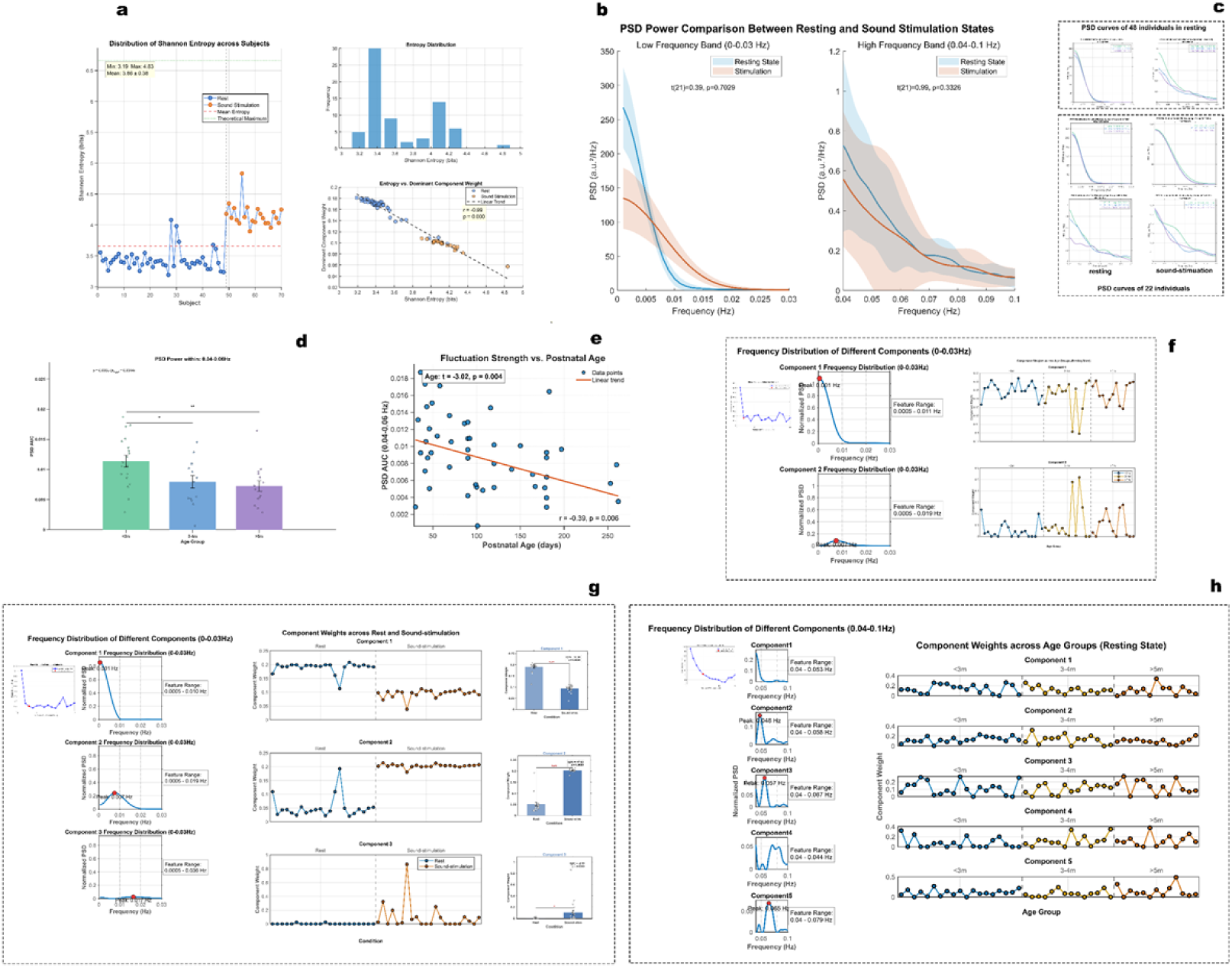
∣ **The fluctuation of clustering coefficient (*C*_p_) in the prefrontal cortex. a**,nPSD complexity analysis of *C*_p_ fluctuations (*n*_rest_=48, *n*_stimuli_=22). **b**, Comparison of resting and sound stimulation PSD AUC in the 0-0.03 Hz and 0.04-0.1 Hz frequency ranges (*n*=22). **c**,Comparison of the PSD curve in the 0-0.03 Hz and 0.04-0.1 Hz frequency ranges across different age groups (*n*_rest_=48, *n*_stimuli_=22). **d**,Comparison of the PSD AUC in the 0.04-0.06 Hz frequency range during resting state (*n*=48) across different age groups (ANOVA; *p* = 0.006;*ω*² = 0.163;95% CI (-0.023, 0.338)). **e**,GLM analysis (*p* = 0.004;*β* = -2.84 × 10⁻⁵;95% CI (-4.74 × 10⁻⁵, -9.47 × 10⁻⁶)) and Pearson correlation analysis (bottom right; *r*=-0.39; *p*=0.006) of the PSD AUC in the 0.04-0.06 Hz frequency range during resting state (*n*=48), related to age. **f**,NMF decomposition of the nPSD in the 0-0.1 Hz frequency range (only showing 0-0.03 Hz) during resting state (*n*=48) using the elbow method to extract 2 components, with comparison of component weights across different age groups. **g**,NMF decomposition of the nPSD using the elbow method to extract 3 components (*n*=44), with comparison of component weights before and after stimulation. **h**,NMF decomposition of the nPSD in the 0.04-0.1 Hz frequency range during resting state (*n*=48) using the elbow method to extract 5 components, with comparison of component weights across different age groups. (*\*p* < 0.01; *\*\*p* < 0.01; *\*\*\*p* < 0.001)

The fluctuations in the clustering coefficient (*C*_p_) in the 0.04-0.06 Hz frequency domain (fig. 4d) showed significant inter-group differences after FDR correction (F (2, 45) = 5.687, *p* = 0.006, *ω* ² = 0.163, 95% CI [-0.023, 0.338]). Notably, although the point estimate suggested a moderate effect, the 95% confidence interval was wide and included zero, indicating that the precision of the effect size estimation based on the current sample size is limited. The effect may exist in the general population but with a small amplitude or may not be stable. We also conducted a GLM analysis on age and gender, and the main effect of age showed a significant linear decreasing trend (fig. 4e) (F (1, 45) = 9.12, *p* = 0.004; *β* = -2.84 × 10⁻⁵, 95% CI [-4.74 × 10⁻⁵, - 9.47 × 10⁻⁶]), while gender had no significant effect. The fluctuations in shortest path length (*L*_p_) did not show significant inter-group differences after FDR correction for two frequency ranges. However, the GLM analysis showed a significant linear decreasing trend for age in the 0.06-0.08 Hz and 0.08-0.1 Hz ranges (fig. 5d-e) (GLM ; *p* = 0.006;*β* = -2.64 × 10⁻⁶;95% CI [-4.49 × 10⁻⁶, -7.87 × 10⁻⁷] . *p* = 0.014;*β* = -1.33 × 10⁻⁶;95% CI [-2.37 × 10⁻⁶, -2.82 × 10⁻⁷]). The fluctuations in global efficiency (*E*_g_) showed significant inter-group differences after FDR correction in the 0.06-0.08 Hz and 0.08-0.1 Hz ranges (fig. 6d) (ANOVA; *p* = 0.006;*ω*² = 0.162;95% CI [-0.024, 0.337] . *p* = 0.016; *ω* ² = 0.129;95% CI [-0.037, 0.302]), and the GLM analysis indicated a significant linear decreasing trend for age (fig. 6e) (GLM ; *p* = 0.006;*β* = - 8.23 × 10⁻⁷; 95% CI [-1.40 × 10⁻⁶, -2.47 × 10⁻⁷] . *p* = 0.016;β = -4.18 × 10⁻⁷;95% CI [- 7.53 × 10⁻⁷, -8.27 × 10⁻⁸]). The fluctuations in local efficiency (*E*_loc_) in the 0.05-0.06 Hz range (fig. 7d, e) (ANOVA; *p* = 0.002;*ω*² = 0.21;95% CI [0.002, 0.384]) (GLM ; *p =* 0.001;*β* = -1.12 × 10⁻⁵; 95% CI [-1.77 × 10⁻⁵, -4.69 × 10⁻⁶]) and small-worldness in the 0.08-0.1 Hz range (fig. 8d, e) (ANOVA; *p* = 0.004; *ω* ² = 0.183;95% CI [-0.013, 0.358]) (GLM ; *p* = 0.010;*β* = -4.19 × 10⁻⁶; 95% CI [-7.32 × 10⁻⁶, -1.05 × 10⁻⁶]) both showed correlations with age.

**Fig. 5.**
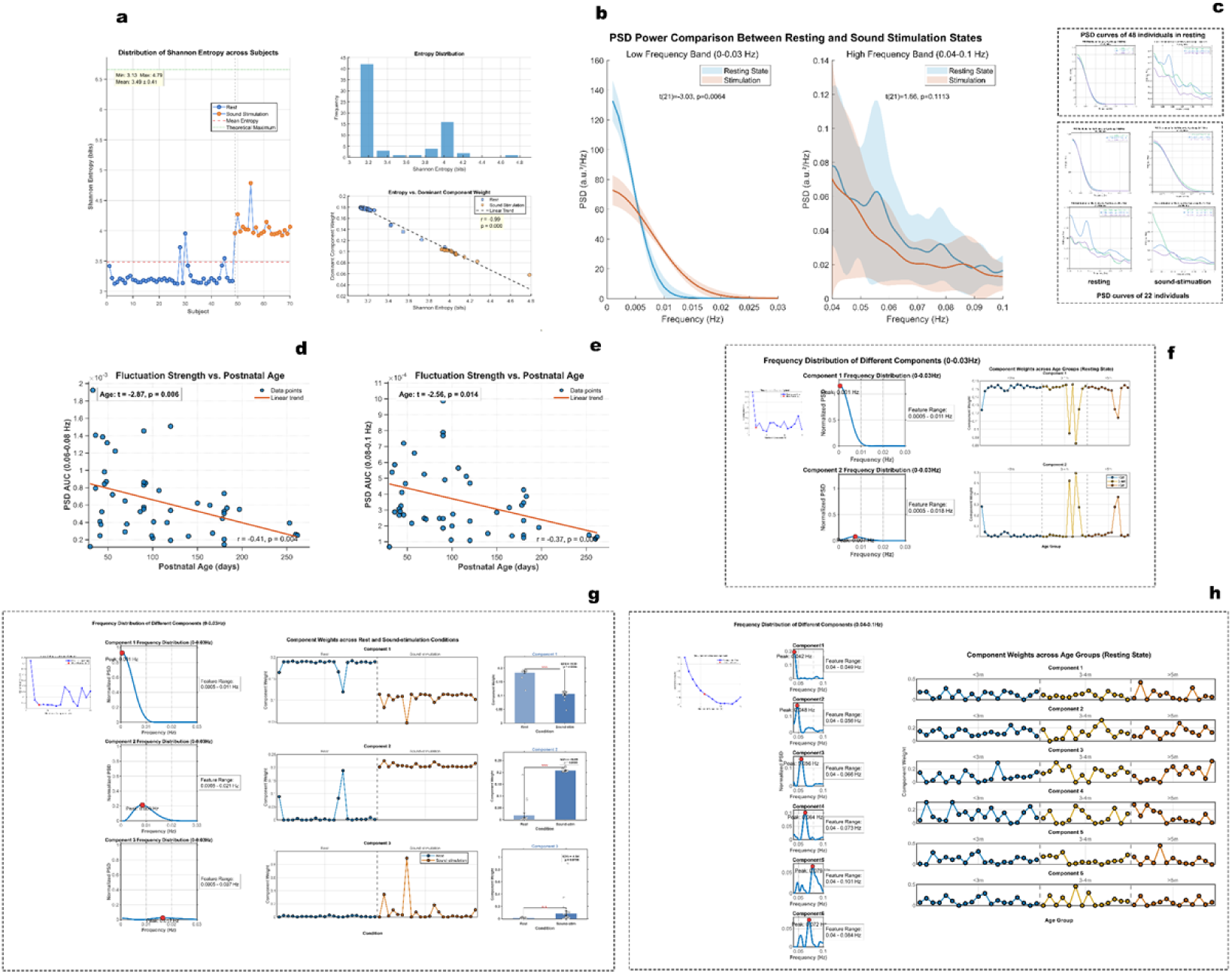
∣ The fluctuation of shortest path length (*L*_p_) in the prefrontal cortex. **a**,nPSD complexity analysis of *L*_p_ fluctuations (*n*_rest_=48, *n*_stimuli_=22). **b**,Comparison of PSD AUC (*n*=22) during resting state and sound stimulation (0-0.03 Hz; *t*=-3.03; *p*=0.006). **c**,Comparison of the PSD curve in the 0-0.03 Hz and 0.04-0.1 Hz frequency ranges across different age groups (*n*_rest_=48, *n*_stimuli_=22). **d**-**e**, GLM analysis and Pearson correlation analysis (bottom right) of the PSD AUC in the 0.06-0.08 Hz (*p* = 0.006;*β* = -2.64 × 10⁻⁶;95% CI (-4.49 × 10⁻⁶, -7.87 × 10⁻⁷)) and 0.08-0.1 Hz (*p* = 0.014;*β* = -1.33 × 10⁻⁶;95% CI (-2.37 × 10⁻⁶, -2.82 × 10⁻⁷)) frequency range during resting state (*n*=48), related to age. **f**,NMF decomposition of the nPSD in the 0-0.1 Hz frequency range (only showing 0-0.03 Hz) during resting state (*n*=48). **g**,NMF decomposition of the nPSD before and after stimulation (*n*=44). **h**,NMF decomposition of the nPSD in the 0.04-0.1 Hz (*n*=48) frequency range. (*\*p* < 0.01; *\*\*p* < 0.01; *\*\*\*p* < 0.001)

**Fig. 6.**
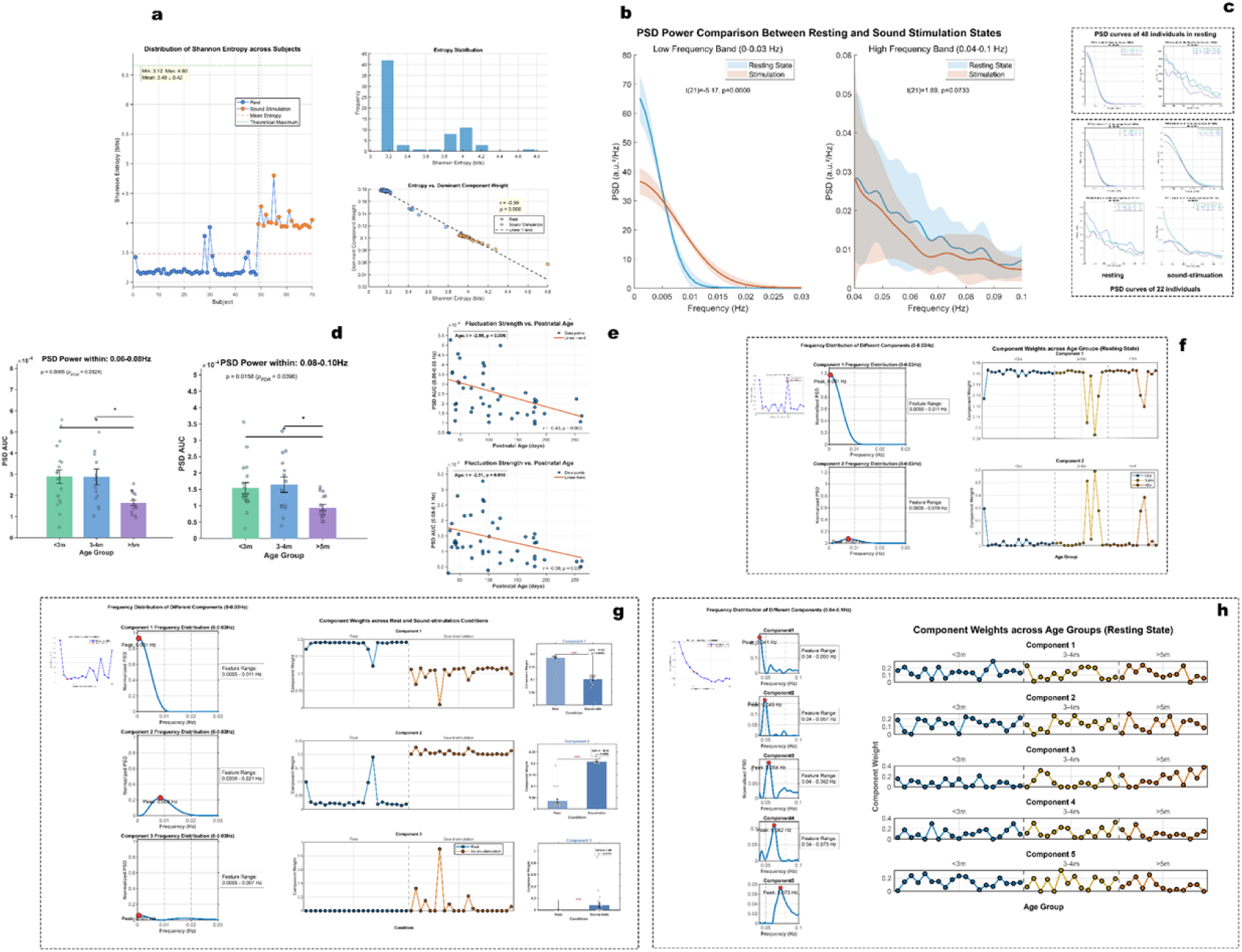
∣ The fluctuation of global efficiency (*E*_g_) in the prefrontal cortex. **a**,nPSD complexity analysis of *E*_g_ fluctuations (*n*_rest_=48, *n*_stimuli_=22). **b**,Comparison of PSD AUC (*n*=22) during resting state and sound stimulation (0-0.03 Hz; *t*=-5.17; *p*=0.00). **c**,Comparison of the PSD curve in the 0-0.03 Hz and 0.04-0.1 Hz frequency ranges across different age groups (*n*_rest_=48, *n*_stimuli_=22). **d**,Comparison of the PSD AUC in the 0.06-0.08 Hz (ANOVA; *p* = 0.006;*ω*² = 0.162;95% CI (-0.024, 0.337)) and 0.08-0.1 Hz (*p* = 0.016;*ω*² = 0.129;95% CI (-0.037, 0.302)) frequency range during resting state (*n*=48) across different age groups. **e**,GLM analysis and Pearson correlation analysis (bottom right) of the PSD AUC in the 0.06-0.08 Hz (*p* = 0.006;*β* = -8.23 × 10⁻⁷; 95% CI (-1.40 × 10⁻⁶, -2.47 × 10⁻⁷)) and 0.08-0.1 Hz (*p* = 0.016;β = -4.18 × 10⁻⁷;95% CI (-7.53 × 10⁻⁷, -8.27 × 10⁻⁸)) frequency range during resting state (*n*=48), related to age. **f**,NMF decomposition of the nPSD in the 0-0.1 Hz frequency range (only showing 0-0.03 Hz) during resting state (*n*=48). **g**,NMF decomposition of the nPSD before and after stimulation (*n*=44). **h**,NMF decomposition of the nPSD in the 0.04-0.1 Hz (*n*=48) frequency range. (*\*p* < 0.01; *\*\*p* < 0.01; *\*\*\*p* < 0.001)

**Fig. 7.**
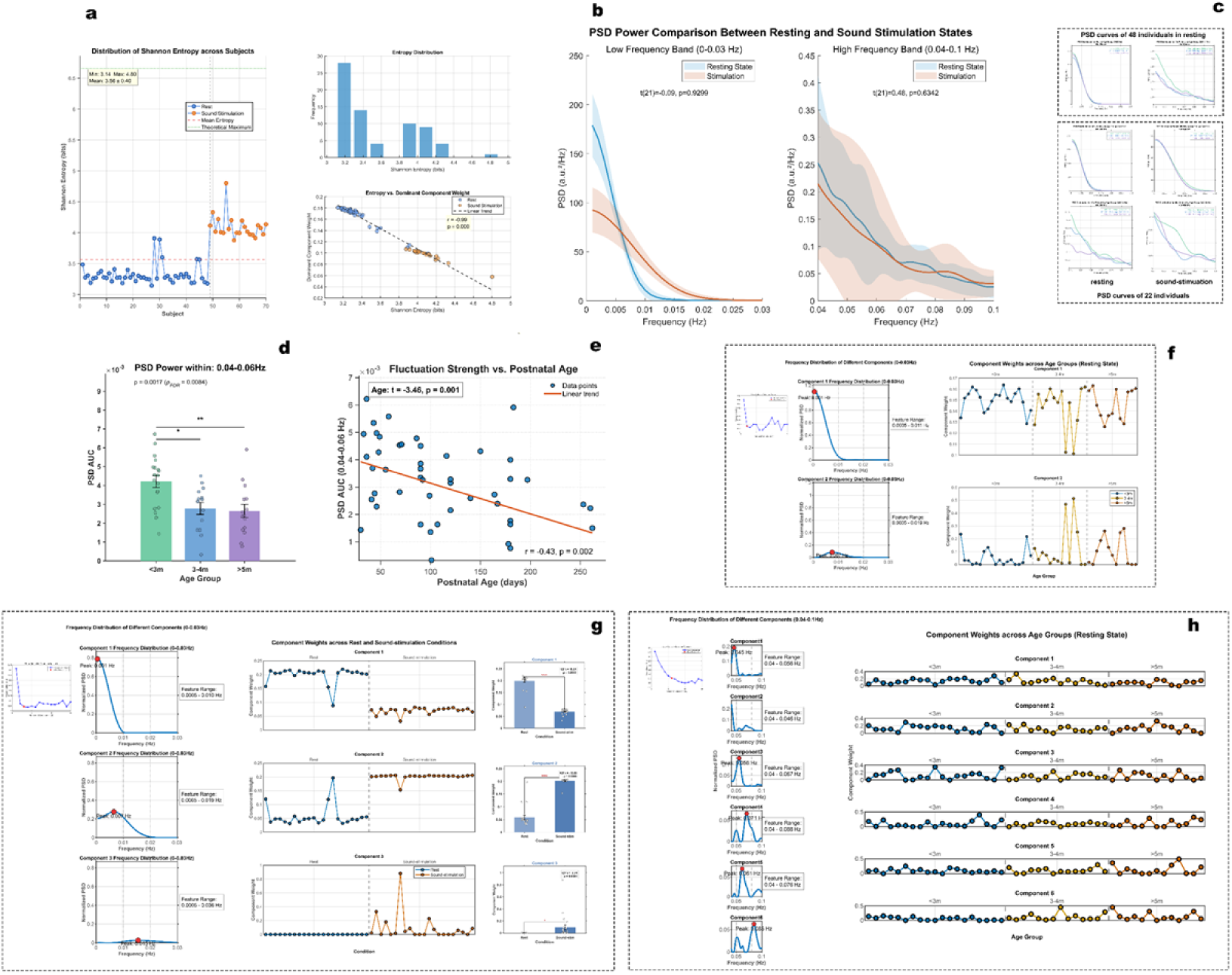
∣ The **fluctuation of local efficiency (*E*_loc_) in the prefrontal cortex. a**,nPSD complexity analysis of *E*_loc_ fluctuations (*n*_rest_=48, *n*_stimuli_=22). **b**,Comparison of PSD AUC (*n*=22) during resting state and sound stimulation. **c**,Comparison of the PSD curve in the 0-0.03 Hz and 0.04-0.1 Hz frequency ranges across different age groups (*n*_rest_=48, *n*_stimuli_=22). **d**,Comparison of the PSD AUC in the 0.04-0.06 Hz (ANOVA; *p* = 0.002;*ω*² = 0.21;95% CI (0.002, 0.384)) frequency range during resting state (*n*=48) across different age groups. **e**,GLM analysis and Pearson correlation analysis (bottom right) of the PSD AUC in the 0.04-0.06 Hz (*p =* 0.001;*β* = -1.12 × 10⁻⁵; 95% CI (-1.77 × 10⁻⁵, -4.69 × 10⁻⁶)) frequency range during resting state (*n*=48), related to age. **f**,NMF decomposition of the nPSD in the 0-0.1 Hz frequency range (only showing 0-0.03 Hz) during resting state (*n*=48). **g**,NMF decomposition of the nPSD before and after stimulation (*n*=44). **h**,NMF decomposition of the nPSD in the 0.04-0.1 Hz (*n*=48) frequency range. (*\*p* < 0.01; *\*\*p* < 0.01; *\*\*\*p* < 0.001)

**Fig. 8.**
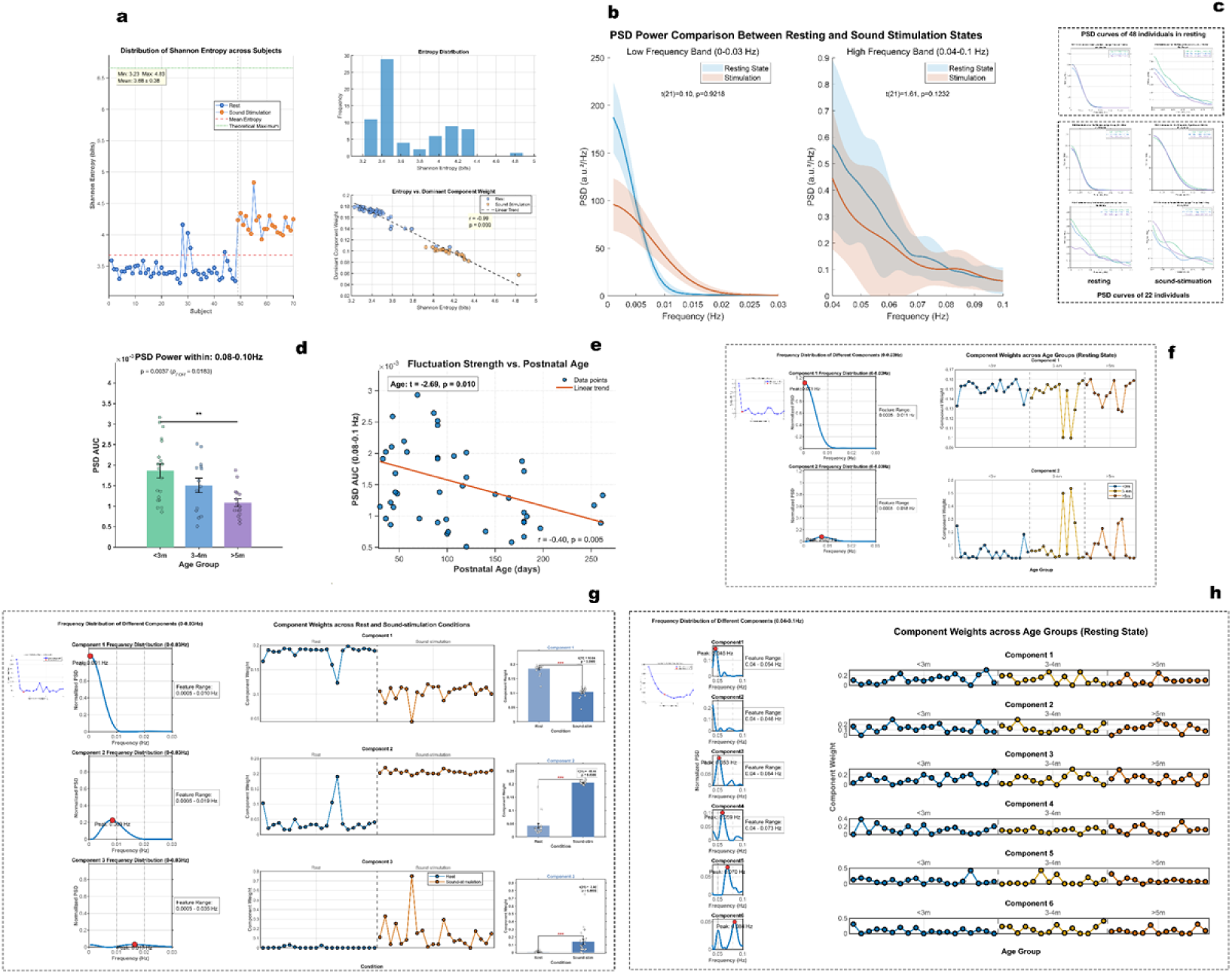
∣ The fluctuation of small-worldness in the prefrontal cortex. **a**,nPSD complexity analysis of small-worldness fluctuations (*n*_rest_=48, *n*_stimuli_=22). **b**,Comparison of PSD AUC (*n*=22) during resting state and sound stimulation. **c**,Comparison of the PSD curve in the 0-0.03 Hz and 0.04-0.1 Hz frequency ranges across different age groups (*n*_rest_=48, *n*_stimuli_=22). **d**,Comparison of the PSD AUC in the 0.08-0.1 Hz (ANOVA; *p* = 0.004;*ω*² = 0.183;95% CI (-0.013, 0.358)) frequency range during resting state (*n*=48) across different age groups. **e**, GLM analysis and Pearson correlation analysis (bottom right) of the PSD AUC in the 0.08-0.1 Hz (*p* = 0.010;*β* = -4.19 × 10⁻⁶; 95% CI (-7.32 × 10⁻⁶, -1.05 × 10⁻⁶)) frequency range during resting state (*n*=48), related to age. **f**,NMF decomposition of the nPSD in the 0-0.1 Hz frequency range (only showing 0-0.03 Hz) during resting state (*n*=48). **g**,NMF decomposition of the nPSD before and after stimulation (*n*=44). **h**,NMF decomposition of the nPSD in the 0.04-0.1 Hz (*n*=48) frequency range. (*\*p* < 0.01; *\*\*p* < 0.01; *\*\*\*p* < 0.001)

The above results indicate that, in the natural sleep state, the fluctuations in prefrontal cortex network properties of 1-8-month-old infants first stabilize in relatively higher frequency bands. The weakening of network properties fluctuations in specific frequency bands represents the gradual maturation of the prefrontal cortex. All the brain network properties observed in this study consistently displayed the same pattern.

### 3.2. Sound stimulation alters the fluctuations in the prefrontal cortex network

To evaluate the change in the complexity of the network fluctuation frequency domain distribution in the sound stimulation state, we calculated the Shannon entropy values of the nPSD for both states. In information theory, Shannon entropy is used as a measure of uncertainty or information complexity (Zhang XD et al., 2017). As shown in the subplots **a** of fig. 3–fig. 8, whether for FC or network properties fluctuations, the task state increased the complexity of the fluctuation frequency domain distribution (For detailed statistics, please refer to the supporting materials: Statistical source data for figures fig. 3-8.a.). That is, under sound stimulation, the power is more evenly distributed across different frequencies, the weight of the dominant frequency decreases, and the brain frequency activity becomes more complex, resulting in increased cognitive load.

By comparing the changes in the PSD intensity before and after sound stimulation in the *n*=22 infants who received stimulation (fig. 3–fig. 8, subplot b), we found that the fluctuation intensity of FC changed little in the large frequency range of 0-0.03 Hz. The fluctuation intensity of network properties, such as *L*_p_ and *E*_g_, generally decreased. In the subplots **b** of fig. 4–fig. 8, we can see that the PSD curves of resting and stimulation states cross at the 0.0055-0.0061 Hz frequency range (Supplementary Fig. 2). A further comparison showed that in the frequency range lower than the crossing point, the fluctuation intensity of network properties in the sound stimulation state decreased, while in the range higher than the crossing point, the fluctuation intensity increased (Supplementary Fig. 2). This demonstrates that the effect of sound stimulation on network properties fluctuations is frequency-specific, leading to a reconfiguration of energy. This crossing effect was not observed in the PSD curve of FC fluctuations (fig. 3b).

The subplots **h, i, j** of fig. 3 and the subplots **g** of fig. 4–fig. 8 show the changes in the principal component weights in the resting and sound stimulation states after performing NMF on the nPSD. (For detailed statistics, please refer to the supporting materials: Statistical source data for figures Fig.3j;Fig.4-8 g)We extracted the common principal components from both the resting and sound stimulation states for the *n*=22 individuals, totaling *n*=44, and compared the differences between the two. All network fluctuations consistently revealed three frequency domain features (fig. 3h, fig. 4–fig. 8g). The first principal component peak is at 0.001 Hz, the second principal component peak is at 0.007-0.009 Hz, and the third principal component generally has two peaks, with the second peak at 0.015-0.017 Hz. From the weight maps generated based on the H matrix, we observed that sound stimulation caused a significant decrease in the first principal component of the network properties fluctuations, a significant increase in the second component, and some network properties saw an increase in the third component (fig. 3i, j; fig. 4–fig. 8g).

### 3.3. Changes in the weight of feature frequency domains are related to the state, not age

We decomposed three stable frequency domain components from all individuals in the resting and sound stimulation states (n=44). The three frequency peaks slightly varied across different networks but were extremely close to each other (fig. 3h, fig. 4–fig. 8g). Whether for FC or network properties fluctuations, we observed significant changes in the weight of these three components across different states (fig. 3i, j; fig. 4–fig. 8g). However, within the same state, the weight of these components did not change with different ages (fig. 4–fig. 8f and h).

Subplot **f** of fig. 4–fig. 8 shows the decomposition of the network properties components in the 0-0.1 Hz range for the resting state, resulting in two principal components. Similarly, the fluctuation of FC in the same resting state decomposed into three principal components (Supplementary Fig. 4), and no age differences were observed. Subplot **h** of fig. 4–fig. 8 shows that when we performed NMF on the relatively higher frequency domain in the resting state, we did not find any correlation between the frequency domain component weights and age.

In addition, when calculating network properties, we also calculated the fluctuation of properties based on Pearson correlation coefficients (Methods), but no frequency domain fluctuations related to age were found. There was only an increase in entropy in the task and stimulation states (Supplementary Fig. 5). When calculating the network properties value for each window based on Pearson correlation coefficients ≥ 0.6, many of the matrix correlation coefficients were too low to compute fluctuations, so we do not recommend using Pearson correlation coefficients as the threshold for calculating network properties. We also calculated the mean, standard deviation, and variance for each pair of channels in the resting state (*n*=48), and only found differences in the standard deviation and variance between groups for one pair of channels (Supplementary Fig. 6).

## 4. Discussion

Exploring the infant brain is more challenging than that of adults because infants cannot cooperate like adults. Most brain resting state research in infants under one year old is conducted during sleep (Agyeman K et al., 2023). The development of certain brain abilities in humans, such as response to name calling, the emergence of joint attention, and the development of self-awareness, all start from infancy. Therefore, studying infant brain development is of great importance, as it encompasses the emergence of numerous cognitive functions from their very beginnings. Investigating these processes may help us understand the formation of intelligence and other critical cognitive pathways. However, given the challenges of cooperation in infants, can we in the future use various stimuli to activate the sleeping infant brain, thereby exploring how the nervous system establishes mature functions? Our research provides an assessment framework based on the maturation of functional connectivity between brain regions for future studies involving stimulation during infant sleep. While our current study has only assessed the prefrontal cortex, in the future we could also evaluate longer-range connections between brain regions. Although preliminary, our study lays the groundwork for future research to explore whether this approach could indicate improvements in connection maturity, thereby reliably distinguishing infants and toddlers with developmental delays and guiding interventions? These questions await further exploration.

Previous research on the development of prefrontal internal connectivity has yielded inconsistent results. Some studies suggest that the internal prefrontal network decreases in the first six months post-birth (Homae F et al., 2010), others report an increase (Gao W et al., 2015), while some show no change (Yin W et al., 2019). These studies have all explored spontaneous fluctuations in sleep states. We believe the inconsistent results stem from individual differences in the prefrontal state of infants during sleep. In our study, we also analyzed whether the strength of functional connectivity and network properties would change with age in continuous signal data. The results showed no significant correlation in resting state, but a marginally significant result in task states (Supplementary Fig. 7). The analysis of the correlation strength of the entire signal misses the detailed temporal information. We believe that the network dynamics of the brain, such as the overall strength of FC fluctuations and the fluctuation characteristics of network properties in specific brain regions, contain more information. Therefore, we explored the frequency domain characteristics of these fluctuations. The results showed that only in the sound stimulation state could FC fluctuations reflect an age-related correlation (fig. 3d, f, g), As shown in fig. 3f, this difference increased with age, showing a clear positive correlation with age. The reason why age differences were only noticeable in the stimulated state could be related to the characteristics of FC. FC represents the degree of synchronization between brain regions (Friston KJ, 2011), and its fluctuations indicate the changes in the strength of synchronization. When stimulation occurs, brain regions associated with the stimulus become activated (Wu YJ et al., 2022), and the functional connectivity strength within specific brain regions increases. This increase in synchronization may become stronger with age (Sun L et al., 2025), leading to more pronounced FC fluctuations. As for why such age-related differences in FC fluctuations were not observed in the resting state, this may be similar to how intelligence differences can be detected in IQ tests but are not as noticeable in everyday life. In the resting state during sleep, there are substantial individual differences in the way individuals are active, which makes it difficult to detect age effects in FC. However, when a stimulus is provided, the brain is placed in a relatively similar state, allowing developmental signs to emerge. This is akin to normalizing the brain’s functional connectivity to reveal differences at various levels. This fluctuation in energy intensity is especially evident in the primary frequency domain we extracted. This indicates that in the sound stimulation state, FC fluctuations in the ultra-low frequency domain strengthen with age.

Network properties, unlike FC, represent the overall properties of connections between things. They are an efficient information processing architecture that strikes a balance between functional specialization and global information integration. We observed that in infants ’ natural sleep states, the fluctuation strength of network properties becomes weaker at higher frequencies, indicating that in the brain’s low-energy sleep state, the maturation of the prefrontal cortex reduces the fluctuations of properties at higher frequencies to conserve energy. Our observation is that this frequency shift occurs above 0.04 Hz (fig. 4–fig. 8d, e), similar to how background apps on a phone close more programs to reduce energy consumption. From the statistical data, we can see that *E*_g_ is the most age-sensitive network properties fluctuation indicator.

Sound stimulation alters the resource reallocation of prefrontal network fluctuations. We found that, in the stimulation state, the weight distribution of fluctuations shifts toward relatively higher frequencies, while the ultra-low frequency domain weight decreases (fig. 3h, i, j; fig. 4–fig. 8g). The energy distribution follows the same changing pattern, with energy reallocated to higher frequencies, showing a distinctly different PSD curve shape (fig. 4–fig. 8b; Supplementary Fig. 2). The principal component weights extracted through NMF show state-dependent stability, and the weights do not increase with age, but the state can alter the weights of the principal components. This result indicates that after sound stimulation, the principal component weights were reallocated. Whether for FC or network properties fluctuations, the reconfiguration rule involves a decrease in the weight of the lowest frequency domain component with a peak at 0.001 Hz, while the weight of higher frequency components increases. Since the nPSD is normalized and energy information from the PSD is removed, the dimensionality reduction analysis purely focuses on the frequency domain morphology, thus avoiding interference from energy levels. Therefore, although no reconfiguration of high and low-frequency energy is observed in fig. 3b, this reallocation of weights is evident in the components. As for why the crossing of the PSD curves in the resting and sound stimulation states is not observed in fig. 3b, we considered that it might be related to age effects. This is because, after directly performing NMF on the PSD, significant age effects were found in both the first and second principal components (Supplementary Fig. 3).

Our findings suggest that during the first six months after birth, internal prefrontal network fluctuations undergo significantly age-related changes. FC fluctuations in the sound stimulation state show significant age-related correlations, while network properties show significance in the resting state. The most age-sensitive network properties indicator is *E*_g_. Auditory stimulation causes a reallocation of prefrontal network fluctuations’ resources, and after stimulation, the weight of higher frequencies increases. The frequency domain weight characteristics of fluctuations do not change with age but change with different states. Overall, our findings are consistent with the view that early prefrontal development may involve a tuning of dynamic network fluctuations toward more optimized patterns, characterized by ’FC enhancing response under stimulation, and network properties improving efficiency at rest.’ This finding provides new mechanistic insights into the functional maturation of the prefrontal cortex and offers potential methods for assessing neurodevelopmental disorders in infants and toddlers in the future.

## 5. Limitations

First, while the total sample size (N=48) is reasonable for infant neuroimaging, the subsample that completed auditory stimulation was relatively small (N=22). Second, the use of a single stimulus (white noise) during natural sleep, though advantageous for control, means our findings are state and stimulus specific. They may not directly generalize to awake states or more complex cognitive tasks. Third, methodological choices inherent to our dynamic connectivity analysis such as the sliding window length and graph theory parameters (e.g., 30% sparsity) could influence the results, although they were based on established practices. Finally, as an observational, cross-sectional study, our findings reveal associations but cannot establish causal relationships between age, brain state, and network fluctuation dynamics.

### CRediT authorship contribution statement

**K.H.L.** Conceptualization, Methodology, Data curation, Software, Writing original draft, editing. **Y.Z.** Resources, Data curation. **Y.H.L.** Supervision.

## Acknowledgments

We thank all of our research participants and their families for their time and participation.

## Data Availability

All relevant data are within the manuscript and its Supporting Information files

## Data Statement

The data that support the findings of this study are available on request from the corresponding authors.

## Appendix A. Supporting information

1. Supplementary information Figs.1-7
2. Statistical source data for figures

## Data availability

All code needed to replicate the results presented in this study has been made available on the Mendeley Data repository at DOI: 10.17632/ts9bgxht4z.1

## Acknowledgements

This work was supported by Research Incubation Fund of Xi’an People’s Hospital (Xi’ an Fourth Hospital) to K.H.L., grant no.CX-37.

